# Study Design: Validation of clinical acceptability of deep-learning-based automated segmentation of organs-at-risk for head-and-neck radiotherapy treatment planning

**DOI:** 10.1101/2021.12.07.21266421

**Authors:** Aman Anand, Chris J. Beltran, Mark D. Brooke, Justine R. Buroker, Todd A. DeWees, Robert L. Foote, Olivia R. Foss, Cían O. Hughes, Ashley E. Hunzeker, J. John Lucido, Megumi Morigami, Douglas J. Moseley, Deanna H. Pafundi, Samir H. Patel, Yojan Patel, Ana K. Ridgway, Erik J. Tryggestad, Megan Z. Wilson, Lihong Xi, Alexei Zverovitch

**Affiliations:** Department of Radiation Oncology, Mayo Clinic, 5881 E. Mayo Blvd, Phoenix, AZ 85054, USA; Department of Radiation Oncology, Mayo Clinic, 4500 San Pablo Rd S, Jacksonville, FL 32224, USA; Google Health, 1600 Amphitheatre Pkwy, Mountain View, CA 94043, USA; Department of Radiation Oncology, Mayo Clinic, 200 1st Street SW, Rochester MN 55902, USA; Department of Quantitative Health Sciences, Mayo Clinic, 5881 E. Mayo Blvd, Phoenix, AZ 85054, USA; Center for Digital Health, Mayo Clinic, 200 1st Street SW, Rochester MN 55902, USA

## Abstract

This document reports the design of a retrospective study to validate the clinical acceptability of a deep-learning-based model for the autosegmentation of organs-at-risk (OARs) for use in radiotherapy treatment planning for head & neck (H&N) cancer patients.

## Introduction

### Background and Rationale

Introduction: An estimated 9.6 million deaths in 2018 were caused by cancer, amounting to one in six deaths globally^1^. Head & Neck (H&N) cancer is among the top six most common cancers in the world^2^. In the United States, H&N cancer accounts for approximately 50,000 of all new cancer cases, and roughly 12,000 deaths in 2010. In 2008, more than 600,000 people were diagnosed with H&N cancer worldwide. Alcohol abuse and long term tobacco consumption are considered to be the major factors in developing H&N cancer, human papillomavirus (HPV) infection also appears to be a risk factor. Moreover, in the United States, there has been an estimated 5% annual increase of oropharyngeal cancers attributable to HPV from the 1980s to 2000s^3^. H&N cancer is a heterogeneous disease, not only due to its aggressive nature, but also because of the complex anatomy in the H&N area.

It is challenging to treat locally advanced H&N cancer with radiation since this modality may lead to damage to salivary glands, mandible, cochlea, larynx, and pharyngeal muscles, which seriously impairs patient quality of life^4^. The use of modern intensity-modulated radiation therapy (IMRT) can help reduce the risk of these complications^5,6^, but this requires the segmentation of the relevant tumor volumes and organs-at-risk (OARs). For H&N cancer, this may require the segmentation of more than 40 structures: manual segmentation for these cases can take up to 3 hours per patient^7^. This can result in delays in the start of patient treatment and limit access to care. It has been shown that provider and institutional experience impacts plan quality and outcomes^8^. Furthermore, a large proportion of H&N cancer patients experience weight loss and/or tumor shrinkage during the course of radiotherapy treatment ^9^. One study in the context of IMRT demonstrated that without treatment replanning, doses to target volumes were reduced in 92% of patients, while the maximum dose to the spinal cord increased in all patients. The study concluded that it is essential to perform repeat CT scanning and replanning to ensure safe radiation doses to healthy tissue whilst delivering adequate doses to target volumes ^10^, for which the time required to perform manual re-contouring is one the biggest barriers to the use of re-planning. The need for H&N replanning tends to be exacerbated in proton therapy and IMPT^11^. Auto-contouring machine learning algorithms can make this process more efficient^12^.

Auto-contouring has been achieved using different techniques, the three main ones being classical image processing^13^ (also known as threshold-based), atlas-based approaches and deep learning. Classical image processing in CT uses algorithms to identify image gradients that distinguish boundaries between, e.g., air filled and water filled tissues or, in case of Positron Emission Tomography (PET), use the metabolic activity between two adjacent tissues; this is based on the Hounsfield Units^14^ in CT and the standardized uptake value (SUV)^15^ in PET. Atlas-based approaches rely on libraries of pre-contoured images as templates to map onto new patient cases using a deformable image registration (DIR) algorithm^16^. Atlas-based approaches have significant overhead in both ascertaining the best match case and modifying the contouring for treatment planning, resulting in only 20% efficiency gain^17^. Deep learning (DL) is a class of machine learning algorithms making use of multilayer neural networks to detect high level features from input data^18^. One study has demonstrated that U-net architecture can produce accurate organ and substructure segmentation^19^. Google Health developed a H&N Segmentation machine learning algorithm (GHHNS V1.0) using 3D U-Net architecture, and demonstrated that GHHNS V1.0 can segment 21 OARs at a human expert level^20^. The DL-produced segmentations were verified by two independent clinical experts, and were measured using surface Dice similarity coefficient (SDSC), which better reflects the task of correcting errors by clinicians than using volumes to quantify deviation. These results showed that the variation SDSC for DL-produced segmentations performed within the threshold defined for human variability in that study for 19 of the 21 OARs^20^.

As part of the Mayo Clinic partnership with Google Health, the department of Radiation Oncology worked with Google Health to improve on the prior work^21^ by Google in automated segmentation. A new Google Health Mayo Clinic DL algorithm (GHMCA) was developed and trained by Google Health using a large cohort of Mayo-derived expert-curated H&N OAR segmentation data. This new H&N model includes 40 organs.

#### Need for a Study

Anatomy contouring plays a crucial part in radiotherapy treatment, as inaccurately defined volumes impact everything downstream, from treatment planning to radiation treatment^22^, which can lead to inferior patient outcome^23^ and decreased patient survival rate^24^. Manual contouring can be time consuming^25^ and can cause delays in treatment planning, which can lead to suboptimal survival rates^26^. With a predicted substantial shortage in radiation oncologists in the US, UK as well as low and middle income countries, it is essential to conduct research towards improving both efficiency and accessibility of high quality patient care.^26,27,28,29^

#### Intended use

This research aims to validate the ability of GHMCA v3b to produce expert level contouring based on Mayo data, and that the radiation dose delivered by radiotherapy plans derived from contouring produced wholly or in part by GHMCA is non-inferior to those derived from human-generated contours in head & neck cancer radiation treatment. If the hypothesis proves to be true, clinical and regulatory experts will be consulted to understand how GHMCA can be integrated into clinical workflows to reduce the cost and time associated with radiation oncology treatment planning, and reduce the time taken between diagnosis and treatment, leading to better outcomes for oncology patients receiving radiation therapy^30.^

## Objectives

### Research hypothesis

The treatment plan time with automated Organs at Risk (OARs) segmentation is significantly lower in H&N radiotherapy compared that with manual physician and dosimetrist contouring alone, without a reduction in quality and accuracy of contouring.

#### Primary Objective

1. To measure time savings in the segmentation of OARs for H&N cancer patients with the use of automatically generated segmentation created using a machine-learning derived model using retrospectively identified radiation therapy planning CT images.

#### Secondary Objectives

2. To investigate the quality of radiation therapy treatment plans derived from OARs generated either wholly or in part by machine learning algorithms as compared to those derived from human generated structures in head and neck cancer treatment, using retrospective radiation therapy planning CT images.
3. To quantitatively investigate the quality and accuracy of OAR contours generated either wholly or in part by machine learning algorithms as compared to those derived from humans in H&N cancer treatment, using retrospective radiation therapy planning CT images.
4. To gather subjective feedback on the human experience of H&N OAR contouring with and without automated segmentation assistance.

## Study Design

A prospective single-blinded (physician verification) randomized mock clinical study was designed to quantify the potential efficiency gains associated with AI-assisted human contouring (Arm 2) in comparison to standard clinical practice of human-only contouring (Arm 1).

For this study, we will recruit and enroll up to 30 Mayo Clinic staff (study participants) that are involved with the routine treatment planning for H&N cancer radiation therapy patients and who possess clinical expertise in H&N OARs segmentation. Study subjects will consist of physicians, certified medical dosimetrists (CMD), and/or medical dosimetry assistants (MDAs). By Mayo Clinic definition, MDAs are professional radiation therapy technicians (RTT) who specialize in tasks associated with treatment planning preparation and adaptive plan evaluation such as OAR segmentation, image fusion and dosimetric plan verification; as such, they can be viewed as proxies for dosimetrists for these specific tasks. The two aforementioned study arms will utilize existing treatment planning CT scans for patients that have previously received treatment. The study participants will be allotted protected time to carry out all study-related tasks and be asked to simulate their nominal clinical mindset while performing these tasks. While the study is prospective in terms of collecting information pertaining to human OAR contouring tasks, the CT scans being utilized are derived from retrospectively-identified H&N patient plans, Therefore, this study will not have any impact on H&N cancer patient treatment.

The CMD/MDA-participants will track time spent on Arm 1 tasks (and stop and restart the timing mechanism during any breaks). At the time of allocation, the physician-participants will be assigned to review OARs segmented on the planning CT by either the CMDs/MDAs c (Arm 1) or by AI (Arm 2). In order to attempt to reduce recall-related bias, a given physician will not contour the same patient for both Arms 1 and 2, but rather, block randomization without replacement is utilized to ensure an equal number of plans per Arm, while eliminating the possibility of evaluating the same patient in both Arms. The physicians will also track time spent performing their tasks (and stop and restart the timing mechanism during any breaks). Each physician-participant will review and revise an equivalent number of cases for both arms, 1 and 2. All self-timed measurements, specifically total time spent for study-related contouring tasks for a given case, will be tracked and entered into an internally-secure, de-identified database for storage and analysis (REDCap Cloud, Encinitas, CA, US). After each physician-participant completes an individual case, an electronic survey for qualitative feedback will be administered using REDCap. Similarly, upon completion of work for all assigned cases, the physician-participant will be unblinded to their previously completed cases and complete an electronic survey evaluating and summarizing their experiences in the study.

To evaluate whether the potential effort reduction introduces clinically-relevant variations in the OAR segmentation, geometric similarity metrics will be computed between the final contours produced in both arms for a given patient study. Furthermore, to assess the difference in the scope revision that the physicians found necessary in each arm, geometric similarity metrics will be computed comparing the individual contours before and after the physicians review and revise the contours for a given patient data-set. The distributions of the geometric similarity metrics for each arm will be compared as discussed in the Outcomes section below.

In addition, the impact on treatment planning will be evaluated using the difference in dose-volume histogram (DVH) metrics calculated for each set of contours, using the same Reference Dose Distribution (RDD) for both arms. For each patient data-set, a clinical-quality volumetric modulated arc therapy (VMAT) plan will be produced by a separate team consisting of an experienced physician, dosimetrist, and medical physicist who will not be participants in this study. This team will produce the treatment plan to meet the same criteria as a plan intended to be used for treatment for H&N cancer. This VMAT planning will be based, in part, on OARs derived from the GHMCA model, wherein the raw OAR model output will be quality-reviewed and potentially revised in a fashion consistent with routine quality assurance for clinical radiation treatment planning at the Mayo Clinic. Examples of applicable minor corrections would be removal of erroneous small-volume segments, i.e., “ditzels,” removal of small-volume holes, or corrections for gross anatomical inaccuracies derived from surgical or disease-related alterations of anatomy. The clinical target volumes derived from the retrospective patients’ treated H&N disease will be copied into the structure set containing finalized OARs; this full set of contours will then be used for optimization structure preparation, per standard clinical practice. Prescription doses will be taken directly from the treated patients’ radiation therapy plans. Thus, the expert CMD and expert MD will generate a clinically viable “reference” VMAT plan per retrospective planning CT, which will undergo patient-specific quality assurance review (plan check and experimental delivery by the expert Ph.D. medical physicists or their proxy) consistent with Mayo Clinic practice. The final reference VMAT plan generated by this process will provide a reference 3D radiation dose distribution (RDD) defined in the same DICOM coordinate frame-of-reference as the retrospective planning CT. Sampling from this singular RDD, DVHs will be generated for 1) expert-AI OARs (that were the basis for the RDD) 2) “gold standard” OARs generated independently by the non-study participant expert MDs and 3)-4) for the OARs generated per each of the two study arms, allowing for an exploratory comparative analysis of DVH-based statistics (including, e.g., a relevant DVH-statistic sensitivity analysis to observed OAR geometric differences). Further VMAT reference planning details are provided in Appendix A, and the evaluation method is outlined in the Outcomes section.

To assess the overall clinical impact of these differences on the treatment planning process, we introduce the normalized Plan Quality Metric (nPQM, see Appendix B). This is adapted from the Plan Quality Metric concept first introduced by Nelms, et al., 2012^31^. This metric is a weighted sum over “scores” assigned to a defined list of treatment planning objectives, where the score is assessed based on how well an individual treatment plan performs on that metric. For this study, all of the scored treatment planning objectives were based on DVH-metrics, as discussed in Appendix B The nPQM accounts for differences in the number of OARs and target volumes by normalizing the combined score to the range [0-100]. This modification is Important for this study (which represents a heterogeneous mix of different H&N cancer histologies, stages and sites), as as to improve combined metric inter-comparability across the set of reference VMAT plans. We propose that, in the present context, such an analysis (involving RDD resampling and use of the nPQM) may serve as an unbiased proxy for elucidating meaningful plan quality issues that could result from OAR geometric differences. We rationalize this approach given that we cannot reliably perform clinical-quality treatment planning per study arm without introducing strong clinician and dosimetrist biases into the generated dose distributions per arm.

## Methods: Participants, Interventions and Outcomes

### Study setting

This non-interventional, prospective study will be conducted at two Mayo Clinic sites, Rochester, Minnesota; Scottsdale/Phoenix, Arizona; using de-identified retrospective H&N radiotherapy planning CT imaging studies collected from Mayo Clinic imaging and treatment planning systems.

The de-identified studies will be passed to the Google research team, where autocontours will be produced using Google’s internal DL tools for non-clinical research use only. Data transfers between Mayo & Google will utilize a secure cloud platform with data encryption in transit and at rest.

### Eligibility criteria

#### Inclusion and exclusion criteria for study participants

##### Inclusion criteria

- Mayo Clinic Department of Radiation Oncology staff from the Midwest and Arizona
- One of the following job roles:
  - Physicians
  - certified medical dosimetrists (CMD)
  - medical dosimetry assistants (MDA)
- Familiarity with the radiation treatment planning of H&N cancer patients in routine clinical care.

##### Exclusion criteria

- Mayo Clinic Department of Radiation Oncology staff (enterprise-wide) involved in algorithm development and/or the design/execution of this validation study.

#### Inclusion and exclusion criteria for de-identified CT dataset

##### Inclusion criteria

- Retrospectively-selected cohort of adult (> 18 years of age) Mayo Clinic patients who had previously given consent to be have their electronic data used for future research purposes and who had radiotherapy planning CT scans prior to radical radiotherapy treatment for H&N cancer treatment performed in the Radiation Oncology departments located in Rochester, MN or Phoenix/Scottsdale, AZ. All tumor types, staging, and histology are eligible for inclusion.
- Considerations were adopted that attempted to balance the cohort demographically in terms of race, diagnosis, biological sex and age.
- CT scans acquired with the intent to be used for radiotherapy treatment planning that had an accompanying high-resolution reconstruction of the acquired data were eligible to be included in the study. If a contrast-enhanced CT was acquired in the same position for the same treatment site during the same imaging session, that CT was also included in the data-set for the given patient.

##### Exclusion criteria

- Patients who are currently undergoing radiotherapy treatment
- Patients who have opted-out of their data being used for research purposes, even in de-identified form,
- Patients who had substantial anatomical changes as a result of previous radiotherapy that an experienced H&N physician determined would cause substantial impact on manual segmentation or radiotherapy planning.
- Insufficient planning CT quality or issues with CT protocol used:
  - CT was not acquired using the institution’s standard protocols for H&N treatment planning
  - The planning CT was deemed to be of inadequate image quality for treatment planning under typical conditions (e.g., motion, swallowing or other significant reconstruction artifacts)
  - A high-resolution (“small field-of-view”) CT reconstruction was not available.
  - Patients with planning CTs with non-standard slice thickness (2mm)
- Contrast-Enhanced CT studies were excluded from the treatment planning CT data-set for a given patient if they were not acquired with a standard protocol or were not of sufficient quality for contouring.

### Interventions

#### The intervention differs in each arm of the study

*In Arm 1:* “Human” arm will entail the participants contouring 40 H&N OAR structures on an de-identified treatment planning CT scan. The CMD/MDA-participants will be instructed to contour as per routine clinical workflow. The physician (MD) will revise these OARs as necessary to meet their clinical expectations.

*In Arm 2:* “AI + Human” arm will entail having the GHMCA automatically produce 40 H&N OAR structures followed by the MD-participants revising and finalizing these OARs as necessary to meet clinical expectations.

A total of 20 de-identified (previously treated) patients’ planning CTs will transit both arms resulting in a total of 40 contour reviews needed by the MDs in total. Each MD participant will perform at least 4 contour reviews (2 cases from each arm), time permitting..

1. All participants will be self-recorded during their contouring work using audio and video screen capture technology with the Mayo Clinic approved Kaltura software. This will allow for time to be measured for each contouring task. In addition, any feedback given verbally during the contouring session can be reviewed for the qualitative feedback portion of the study. Video recording will not be reviewed until the time of analysis.
2. The sequence of approaching Arm 1 or Arm 2 first for the physicians will be randomized for each de-identified patient case. A different physician will be assigned to each arm for each case to minimize bias. Each physician will need to complete an equivalent amount of Arm 1 and Arm 2 cases.
3. After work is completed for all allocated patient cases, each participant will be approached by a member of the Mayo Clinic research team for subjective feedback in the form of an electronic survey.

#### Algorithm Version Used In Study

GHMCA v3b

#### Details on Input Data Acquisition and Selection

The input data for the AI-model used in this study will be selected from previously acquired data-sets from patients who meet the eligibility criteria defined above, and then inspected to ensure that the CT series associated with the planning of their H&N radiation therapy meet the specified imaging-series eligibility criteria. From this pool of acceptable CT studies, the data for 20 patients will be selected in order to balance the cohort demographically in terms of race, diagnosis, biological sex and age, as well as to have the cohort reflect the departments’ treatment demographics in terms of tumour types, staging, and histology. In a future manuscript to be prepared pertaining to the study findings, the demographics of the patients whose planning CT datasets were included will be summarized.

#### Human-AI Interaction in Handling of Input Data

There is no Human-AI interaction as part of the AI-intervention.

#### Output of AI Intervention

For each set of retrospectively acquired (and de-identified) imaging data for a single patient, the AI-algorithm will produce a set of volumetric segmentations of up to 40 individual OAR structures registered to the treatment planning CT. These segments will be formatted into a Digital Imaging and Communications in Medicine standard-Radiotherapy Structure set (DICOM-RS)^32^ file for transfer to the treatment planning system.

#### Contribution of AI-Intervention to Clinical Practice

This is an *in silico* investigation of the impact of the AI Intervention on the clinical workflow using retrospectively acquired (and de-identified) data, with no impact on the patient’s course of care. There is no contribution of the AI-Intervention to any clinical practice related to this study.

#### Criteria for Discontinuing or Modifying Intervention For Given Participant

If a participant opts to withdraw from the study or otherwise not completed the allocated tasks, the data associated with that participants activities and feedback will not be reviewed or included in the analysis. An additional participant will be recruited to perform the tasks that had been assigned to the withdrawn participant.

#### Strategies to improve Adherence to Intervention Protocols

Prior to performing any activities associated with the study, the participant will meet with a study coordinator to receive training and the participant will be provided with documentation that explicitly states the expectations for completing the activities and detailed instructions for how to perform the activity. They will also be given contact information for a point of contact to address questions about the study or technical issue encountered. The participants will receive additional protected research time to perform the activities associated with this study, and specific time will be scheduled for these activities that will be blocked from conflicting appointments.

### Outcomes

#### Primary outcome

The primary hypothesis of this study is that the use of AI-assistance (Arm 1) will result in at least a 30% reduction in the total time required by participants for the contouring process, compared to contouring without AI-assistance (Arm 2), which represents the current standard clinical practice. Total time from assignment until completion of verification will be recorded and the percentage reduction from Arm 1 will be calculated for each of the 20 assigned plans.

#### Secondary outcome

##### Qualitative Analysis

Survey results will be reported using descriptive statistics. Interview data will be analyzed thematically.

##### Quantitative Analysis

We will quantitatively assess the potential impact of the use of the AI-generated contours had they been used as part of the clinical treatment planning process. The statistical methods employed for the following comparisons are described in the Statistical Analysis section.

1. The mean geometric similarity metrics (Dice Similarity Coefficient, Hausdorff Distance, Added Path Length) of the OAR structures will be compared before and after physician revisions between the two arms.
2. Comparisons of the minimum and mean absolute doses received by each structure, and the minimum absolute dose received by the sub-volume of 0.03cc receiving the highest doses of the structure (D0.03cc)^33^, for each structure will be compared with the doses received by the same structure created in the other arm for the same patient data-set, using the RDD described in Appendix A.
3. A comparison of the change in the normalized Plan Quality Metric (nPQM, described in Appendix B) calculated for each case using the structures in each arm relative to the nPQM resulting from the expert-AI OARs, computed using the RDD.
4. Comparisons of differences in geometric similarity measures between the individual organ-at-risk segmentations between the two arms (Dice Similarity Coefficient, Hausdorff Distance, Added Path Length and any other measures applicable).

### Participant timeline

After the potential participants have indicated interest, time will be scheduled for them to meet with study staff to discuss the study and to provide consent. If consent is given, the study staff will then provide initial training and provide documentation for review. Research participants will be provided protected time to perform allocated activities in the study to the amount of 4 hours for each CT scan contoured. Electronic subjective experience questionnaires will be administered after each case reviewed by MDs. If further clarification or detail is required, interviews will be scheduled after all cases have been reviewed.

### Sample size

There will be 20 assigned plans in each arm. Estimates based on previously curated, internally collected, data on timing results for manual contouring without auto-contouring (Arm 1) compared to with auto-contouring assisted manual contouring (Arm 2) will be utilized to power this study’s primary hypothesis. This data demonstrated an average reduction in time spent of approximately 65% with a standard deviation of reduction of 20%. We will utilize a one-sided paired t-test based on proportion of time reduced by utilizing AI-assisted manual contouring (Arm 2). We hypothesize that, in order to be clinically impactful, we need to see at least a 30% reduction in time spent contouring based on fully manual contouring (Arm 1). We will conservatively estimate, based on prior data, that we achieve at least a 50% reduction in time spent contouring. Given these estimates and utilizing an **α** = 0.025 we will have approximately 92.4% power to show that the average time reduction is significantly greater than 30%.

### Recruitment

A recruitment email will be sent to all radiation oncologist in the participating departments who meet the criteria to participate in the study, including an overview of the purpose of the study, the activities they will be asked to perform and how much time their participation will require, and a notification that they will have the allotted time dedicated to their participation. A similar recruitment email will be sent to the CMD and MPA groups within the participating departments.

## Methods: assignment of interventions

### Sequence generation

The sequence will be generated using block randomization relying on computer-generated random numbers.

### Allocation concealment mechanism

The participant radiation oncologists will have the origin of their allocated contours concealed. They will not be informed whether the contours were generated by the AI-model, a participant dosimetrist or dosimetrist’s assistant. To ensure concealment, the data management will be performed outside of the clinical treatment planning system that will be used by the participants in the study. The data-sets will be de-identified prior to import into the treatment planning system, and all objects specific to the arm of the intervention (such as the object’s Id or Name fields) will be labeled with the participant radiation oncologists’ initials. No mention of the allocation will be made in the notification materials given to the participants indicating which cases they have been allocated, and the study staff directly interacting with the participants will also have the allocation concealed to them. After the radiation oncologist participants finish reviewing and revising all allocated cases, and the participants have completed the assigned surveys for those cases, they may have an interview scheduled, in which case the allocation will be revealed to get feedback on the experience.

### Implementation

Block Randomization without replacement will be utilized in order to assign de-identified, blinded contour sets to physician-participants. Randomization was completed utilizing R (ver 4.0.3) and implemented in order to ensure each participant will evaluate an equivalent number of plans per arm, while reducing possibility of recall bias by ensuring they do not contour plans in both arms from the same patient. The statistical team will complete the randomization and enter the randomization into the REDCap database, thereby blinding the process to the rest of the study team. Participants will be consented and enrolled by Mayo Clinic Radiation Oncology Clinical Research Coordinators. Once enrolled and consented the statistical team will utilize the REDCap database, and previously described randomization algorithm, to electronically assign de-identified, blinded contour sets to the participant. All subsequent surveys, assignment of new plans, and study materials will be electronically assigned.

### Blinding (masking)

The physician-participants will not be informed of the origin of the contours (auto-segmentation by the DL-model or manually generated by the CMD/MDA-participant) that they have been asked to review until after they have completed the contouring of all allocated cases and the completed all of the case-specific participant surveys. In follow-up interviews after completion of these tasks, the participant-physician may be unblinded as to the origin of the specific cases that they were allocated to assess differences in perception of the quality of the initial contours or the burden required by the revision task.

## Methods: data collection, management and analysis

### Data collection methods

To ensure that the participants have a consistent experience during the study, the on-boarding documentation is the same for all participants within one job category (physicians or dosimetry staff), which is provided with this document. Standardized instruction was provided to the study coordinator staff to ensure uniform interaction with the participants. For the study personnel involved in the generation of the plan used to calculate the Reference Dose Distribution (RDD), uniform instructions were provided as detailed in relevant sections of this document.

### Data management

All study data will be stored electronically in specific applications (REDCap) managed by the Mayo Clinic information technology, behind the Mayo Clinic firewall, and security teams and with access limited by the institutional user right’s management policies and procedures, except when a copy of the data is transferred securely to Google Health for the AI-intervention (which will then be managed by the Google Health team). Redundant copies of all treatment planning-related files (images, structure sets, treatment plans, radiation doses) will be stored in different databases. Prior to analysis of the survey, geometric, or dosimetric data, the data integrity will be verified as discussed in the auditing section below, as well as after the transfer of any of the treatment-planning related data between systems or databases. If any unanticipated discrepancies are discovered at any of these steps, a select team of study investigators responsible for data management will be informed, and they will make a recommendation to the principal investigators approval for how to resolve the issue.

### Statistical methods

#### Primary outcome

The primary outcome is designed as a one-sided paired t-test based on proportion of time reduced by utilizing AI-assisted manual contouring (Arm 2). The statistical test will be conducted to ensure that the average time saving is significantly greater than 30%. Therefore, we will consider the study to be successful and worth further study given statistical significance (p<0.05) or the lower bound of a one-sided confidence interval greater than a 30% reduction.

#### Secondary outcome

##### Qualitative Analysis

Survey results and interview responses will be collated and descriptive statistics will be provided independently for all questions individually as well as composite scores provided for the User Experience questionnaires based on the NASA TLX survey^34,35^. When appropriate nonparametric Wilcoxon Rank Sum and Fisher’s Exact Tests may be utilized and statistical significance may be evaluated with two-sided tests with an **α** = 0.05.

##### Quantitative Analysis

We will quantitatively assess the potential impact of the use of the AI-generated contours had they been used as part of the clinical treatment planning process. We will present descriptive statistics and 95% confidence intervals (95%-CI) for all endpoints. Corrections for multiple testing in order to control the type I error rate will be implemented in order to ensure appropriate conclusions. Specific considerations for each aim are listed below:

1. Comparisons utilizing two-sided paired t-tests will be performed to determine if there are significant differences between the mean geometric similarity metrics (Dice Similarity Coefficient, Hausdorff Distance, Added Path Length) of the OAR structures before and after physician revisions between the two arms.
2. Dosimetric comparisons utilizing two-sided paired t-tests will be performed to determine if there are significant differences between the individual structures in each arms in terms of the minimum absolute dose received by the sub-volume of 0.03cc receiving the highest doses of the structure (D0.03cc)^33^, and the mean and minimum doses of the structures based on a common reference dose distribution (RDD) generated as described in Appendix A.
3. A comparison using a two-sided paired t-test will be performed to determine if there is a significant difference between the change in the normalized Plan Quality Metric (nPQM, described in Appendix B) calculated for each case using the structures in each arm relative to the nPQM resulting from the expert-AI OARs, computed using the RDD.
4. Comparisons utilizing two-sided paired t-tests to determine if there are significant differences in geometric similarity measures between the individual organ-at-risk segmentations between the two arms (Dice Similarity Coefficient, Hausdorff Distance, Added Path Length and any other measures applicable).

## Methods: monitoring

### Data monitoring And harms

There will be no formal data monitoring committee: there is no intervention to patient care associated with this study and no interim analysis will be performed nor any evaluation of harms. However, throughout the study the statistical team will monitor physician-participant protocol adherence, as well as, completion rates for survey questions in order to ensure complete data.

### Ethics and dissemination

#### Research ethics approval

This study has been approved by the Mayo Clinic Institutional Review Board (Study no: 21-008372) on 11/2/2021.

#### Protocol amendments

None

#### Consent or Assent

##### Radiotherapy Patients

After approval from the Mayo Clinic Institutional Review Board, 20 patients were identified as meeting eligibility criteria. All patients were confirmed to have provided consent for their data to be used in future research.

##### Mayo Clinic Staff Participants

Study coordinators will meet with potential participants virtually to obtain written consent, including informing the potential participant that their participation is volunteer and will not impact their employment in any way. The participants will use electronic consent technology or a signed paper consent at the choice of the participant. The documentation will be stored electronically and may be printed as well. The subject may contact the study team to provide a copy of the signed form.

### Confidentiality

All study data with patient protected health information is secured within the institutional computer network at Mayo Clinic. Records regarding the use of a patient’s data in this study will be stored in a database with access limited to study personnel. Before the patient’s imaging data was used in the study, it was transformed at the DICOM data level to meet the HIPAA de-identification standard using the Safe Harbor Method. Furthermore, this de-identification process underwent external review. The de-identified data will be transferred securely to Google Health for the generation of the AI-output. The AI-output data will be transferred securely to Mayo Clinic, and will remain de-identified permanently. All transfers are done using the HITRUST certified Google Cloud Platform (GCP).

The surveys generated by the Mayo Clinic staff participants will be stored in the study management software database by record number without any identifiers for the participant. An electronic document revealing the staff member associated with the record number will be kept separate and only accessible to select study personnel for use in resolving discrepancies in the data and only at the approval of the principal investigator.

## Data Availability

This is a statement of study design, no data is available.

## Declaration of interests

None

## Access to data

The data will not be available for public access because of patient privacy concerns, but are available from the corresponding author on reasonable request. The AI-model and its code will not be made available due to contractual, licensing, and/or commercial restrictions.

## Dissemination policy

The principal investigators will review all manuscripts related to this study for appropriateness and scientific merit prior to submission for publication. This document, and its amendments, represents the full protocol, which will be publicly available including clear documentation of the record and dates of all amendments and modifications. The primary outcome paper of this study will report the primary and secondary outcomes listed in this document. Every attempt will be made to reduce to an absolute minimum the interval between the completion of data collection and the release of study results via publication in an appropriate journal. Other papers, abstracts and presentations must be approved by the principal investigators prior to submission. The study results will be released to the study personnel, the department leadership, and the general medical community.

## Auditing

### Data Transfer Quality Assurance

After any data-transfer of imaging or treatment planning data between different systems or databases, the integrity of the data transfer will be verified prior to use in the new system by the investigators. Any discrepancies will be referred to specific study investigators responsible for data management for resolution.

### Participant Timing Quality Assurance

Prior to the analysis of the screen-capture videos for timing data, study staff will review the videos for accuracy of timing. This will include reviewing whether the videos were started and stopped at the appropriate times (for instance, that they were not left running after the participant completed the study activities or started after the participant had begun doing the activities). It will also be reviewed to see if the participant was not interacting with the computer for an extended period of time, defined as any interval of greater than 2 minutes for which the mouse was not moved. Any excess time will be deducted from the total time for the participant’s activity for the allocated case. If this audit indicates that part of the participant’s activity was not captured by the video, specific study investigators responsible for data management will query the treatment planning data-base for time-stamps and will try to reconcile any discrepancies. If it is necessary to query the data-base to reconcile the missing data, the number of cases will be reported in the primary outcomes publication, and any other study-related publications that make use of the participant timing data in the results sections, and specific information regarding the case will be reported (in either the results section or as supplemental data), including which arm it belongs for, and the total time for the case calculated using both methodologies.

## Appendix A

### Creation of the Reference Dose Distribution

For each individual CT data-set, a volumetric modulated-arc therapy (VMAT) reference plan will be produced to create the Reference Dose Distribution (RDD), which will allow for the evaluation of the impact of the OAR contouring on the dose-volume statistics. The radiation oncology physicians, CMDs, and MDAs involved in the generation of the Reference Plan will all be experienced with planning radiation therapy treatment for H&N cancer, and will not be eligible to participate in the study.

#### Target Volumes

For the reference plan to be created for this study, the same clinical target volume(s) (CTVs) that had been used for the initial treatment will be used. There could be up to 3 different CTVs for a given data-set, high-, intermediate-, and low-risk targets; each target will be prescribed a different dose. Each CTV includes all CTVs with higher risk, and planning target volumes (PTV) will be created from each individual CTV by adding a 3mm expansion (with the PTVs cropped to the patient’s body contour afterward).

#### Organs-At-Risk (OAR)s

The AI-model’s outputs will serve as the organs-at-risk contours used for the creation of the Reference Plan. Before the planning begins, an experienced member of the dosimetry staff (either a CMD or MDA) will review the contours to verify that they have no gross defects, and will perform minor modifications if necessary using the clinical treatment planning system (Eclipse version 15.6, Varian Medical Systems, Palo Alto, CA). To ensure that these modifications are minor, the staff member will be instructed to complete the task in less than 30 minutes, and given a specific list of modifications that they can make. These potential modifications are limited to:

- Conventional post-processing:
  - Removal of isolated contour segments with a volume less than 0.1cc,
  - Regions of volume less than 0.1cc that are not included in a given contour that completely surrounds the region will be included in the surrounding region
- For regions of overlap between distinct OAR structures, the region will be assigned to one structure and removed from the other structure as deemed most appropriate by the staff member performing the modifications.

If the task takes more than 30 minutes, the contours will be restored to their state before the modifications were made and a second CMD or MDA will be asked to perform the review and revision. If this task would again take more than 30 minutes, the physician involved in the generation of the Reference Plan will review the cases and determine if they are acceptable for clinical treatment without modifications. If so, then the un-modified AI-model output will be used. If not, then the case will be excluded from dosimetric analysis.

#### Treatment Prescription

The treatment prescription that will be used to generate the Reference Plan will correspond to the institutional standard for a standard fractionation radiotherapy treatment given each patient’s disease etiology and staging. Standard fractionation in this context means that the patient receives between 25 and 35 daily treatments (5 treatments per week), and at least 1.8 Gy prescribed per fraction to the highest-risk PTV. For cases in which the clinical treatment plan was standard fraction, the same prescription will be applied to the reference case. For patients who were treated using a hypofractionated regime, the equivalent standard fractionation dose (according to the institutional standards) will be used instead. If the patient has multiple target volumes receiving different prescribed doses, the higher dose prescriptions will be achieved using a simultaneous integrated boost.

#### Treatment Plan Optimization

The reference plans will be optimized using the institutional standard beam geometry for H&N cancer treatment planning: 3 full-arc co-planar VMAT beams with a single isocenter, using 6 MV photon beams (with flattening filter). The treatment optimization will be done using Eclipse version 15.6 (Varian Medical Systems, Palo Alto, CA) by a CMD, and evaluated by a radiation oncologist to determine if it would be acceptable to use for treatment as if the Reference Plan has been created for clinical use, using institutional standards and taking into account the specifics of the case under review. The institutional standard treatment planning template for a generic standard-fractionation H&N cancer therapy is shown below. The Reference Plan planning process will continue until the physician deems the case ready for treatment.

## Appendix B

### Normalized Plan Quality Metric

The Plan Quality Metric (PQM) framework is a tool that was developed to create a standardized method for evaluating how well a specific treatment plan satisfies multiple planning objectives^31,33^. The planning objectives often involve specific dose-volume constraints. For each objective, a method must be defined that gives a score for how well a given plan achieves the objective. For instance, keeping the maximum dose of an organ-at-risk (OAR) below a certain threshold may warrant receiving a score of 5 points, with no points awarded if the threshold is not met. Alternatively, the points may be awarded on a sliding scale if it falls within a certain range. When a plan is evaluated using the PQM scorecard, all of the points are tallied to generate the plan’s score. Typically, a PQM scorecard is developed for a specific context, like a study of planning variation over a cohort of patients for a disease site. The form of the PQM should reflect the clinical decision-making within that context.

In this study, an individual data-set may have between one and three planning target volumes (PTVs) and some OARs may have been removed during prior surgery. This means that there will be variation in the number of structures being evaluated, and therefore in the number of criteria available to contribute points to the final PQM score. To allow meaningful comparisons across our study cohort, we will use the normalized PQM (nPQM) score, which divides the PQM score by the maximum score possible given the contours present in the scan (PQM_max_), and then scales to a maximum of 100:

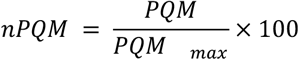

The PQM scorecard to be used for analysis in this trial is shown in Table 1. This consists only of dose-volume objectives. There are two types of functions used to determine the score for a specific objective. The threshold scores give the maximum number of points for that objective if the plan meets the objective and no points if the objective is not met. The linear scores have two thresholds. If the plan achieves the “ideal” threshold for the constraint, the maximum points are given, and no points are given if the plan does not meet the “minimally acceptable threshold”. If the objective falls between the two thresholds, the number of points awarded is determined by linearly interpolating between the two thresholds using the value of the dose-volume statistic.

**Table 1:** Plan Quality Metric for Evaluation of Treatment Plans.

| Structure | Constraint | Function | Thresholds | Max Score |
| --- | --- | --- | --- | --- |
| brachial_plex_l | D0.01cc[Gy] | threshold | < 63 Gy | 5 |
| brachial_plex_r | D0.01cc[Gy] | threshold | < 63 Gy | 5 |
| brain | D0.01cc[Gy] | threshold | < 56 Gy | 5 |
| brain_stem | D0.01cc[Gy] | threshold | < 54Gy | 5 |
| cochlea_l | Mean[Gy] | threshold | < 35 Gy | 1 |
| cochlea_r | Mean[Gy] | threshold | < 35 Gy | 1 |
| constrictors_p | Mean[Gy] | linear | < 59 Gy, < 49 Gy | 2.5 |
| cord | D0.01cc[Gy] | threshold | < 45 Gy | 5 |
| crico_p_inlet | Mean[Gy] | linear | < 59 Gy, < 49 Gy | 2 |
| ctv_high | D95%[%] | linear | > 95%, > 100% | 2.5 |
| ctv_high | V100%[%] | linear | > 90%, > 98% | 2.5 |
| ctv_intermediate | D95%[%] | linear | > 95%, > 100% | 2 |
| ctv_intermediate | V100%[%] | linear | > 90%, > 98% | 2 |
| ctv_low | D95%[%] | linear | > 95%, > 100% | 1 |
| ctv_low | V100%[%] | linear | > 90%, > 98% | 1 |
| **esophagus** | Mean[Gy] | linear | < 39 Gy, < 29 Gy | 2.5 |
| **esophagus_cerv** | Mean[Gy] | linear | < 39 Gy, < 29 Gy | 2.5 |
| **ext_aud_canal_l** | D0.1cc[Gy] | linear | < 60 Gy | 1 |
| **ext_aud_canal_r** | D0.1cc[Gy] | linear | < 60 Gy | 1 |
| **eye_l** | V50Gy[cc] | threshold | < 0.01 cc | 1 |
| **eye_r** | V50Gy[cc] | threshold | < 0.01 cc | 1 |
| **lacrimal_l** | D0.03cc[Gy] | threshold | < 30 Gy | 1 |
| **lacrimal_r** | D0.03cc[Gy] | threshold | < 30 Gy | 1 |
| **larynx-ptv** | Mean[Gy] | linear | < 25 Gy, < 15 Gy | 3 |
| **lung_total** | Mean[Gy] | threshold | < 20 Gy | 1 |
| **mandible** | D0.03cc[Gy] | linear | < 75 Gy, < 67.5 Gy | 1 |
| **nasal_cavity-ptv** | Mean[Gy] | linear | < 45 Gy, < 35 Gy | 5 |
| **optic_chiasm** | D0.01cc[Gy] | threshold | < 50 Gy | 5 |
| **optic_nrv_l** | D0.01cc[Gy] | threshold | < 50 Gy | 5 |
| **optic_nrv_r** | D0.01cc[Gy] | threshold | < 50 Gy | 5 |
| **oral_cavity-ptv** | Mean[Gy] | linear | < 30 Gy, < 20 Gy | 5 |
| **parotid_l-ptv** | Mean[Gy] | linear | < 30 Gy, < 20 Gy | 2.5 |
| **parotid_r-ptv** | Mean[Gy] | linear | < 30 Gy, < 20 Gy | 2.5 |
| **pituitary** | Mean[Gy] | linear | < 30 Gy, < 10 Gy | 1 |
| **ptv_high** | D1[%] | linear | < 112%, < 108% | 5 |
| **ptv_high** | D95[%] | linear | > 95%, > 100% | 5 |
| **ptv_high** | V100[%] | linear | > 90%, > 98% | 5 |
| **ptv_intermediate** | D95[%] | linear | > 95%, > 100% | 2.5 |
| **ptv_intermediate** | V100[%] | linear | > 90%, > 98% | 2.5 |
| **ptv_low** | D95[%] | Linear | > 95%, > 100% | 1 |
| **ptv_low** | V100[%] | Linear | > 90%, > 98% | 1 |
| **submand_l-ptv** | Mean[Gy] | linear | < 45 Gy, < 30 Gy | 2.5 |
| **submand_r-ptv** | Mean[Gy] | linear | < 45 Gy, < 30 Gy | 2.5 |

---

**Figure 1.**
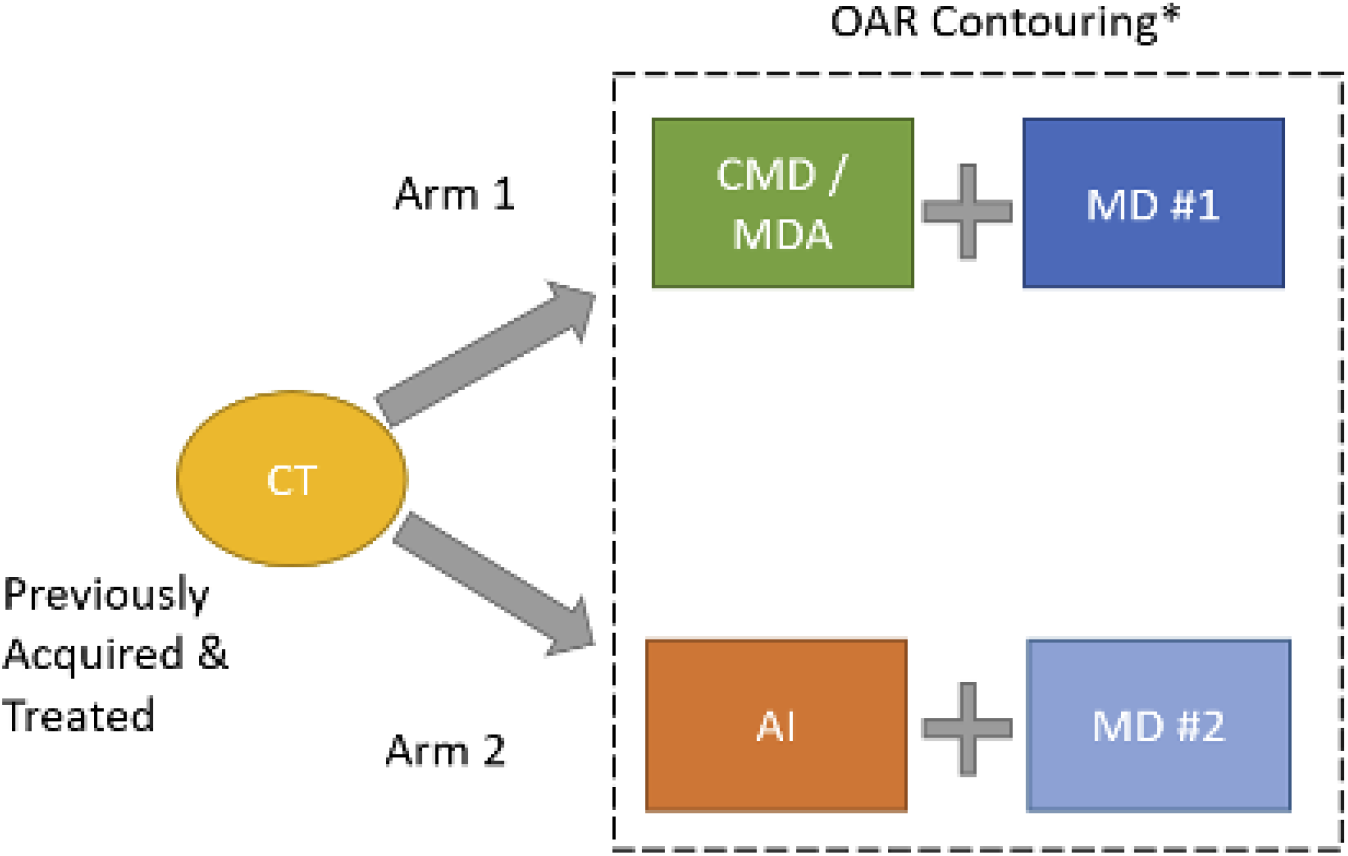
Trial Design Flow Chart

## Notes

### Competing Interest Statement

The authors have declared no competing interest.

### Funding Statement

Mayo Clinic will be responsible for supplying de-identified training data for deep-
learning models. Google Health is responsible for undertaking machine learning research. Mayo and Google Health willbear their own costs in connection with this study.

### Author Declarations

IRB of Mayo Clinic gave ethical approval for this work.

